# Improved mTBI outcomes with a BCI Amplified CRT Training: A retrospective chart review

**DOI:** 10.1101/2020.09.10.20192237

**Authors:** C.T. Cripe, P. Mikulecky, Rebecca Cooper, T. Eagan

**Author notes:** Corresponding author contact info: Peter Mikulecky -.

## Abstract

This study is a retrospective chart review of 200 clients who participated in a non-verbal restorative Cognitive Remediation Training (*r*CRT) program. The program was applied to effect proper neural functional remodeling needed to support resilient, flexible and adaptable behaviors after encountering a mild to medium closed head traumatic brain injury (mTBI). The *r* CRT training program focused on improving functional performance in executive cognitive control networks as defined by fMRI studies. All *r*CRT training activities were delivered in a semi-game-like manner, incorporating a Brain Computer Interface (BCI) that provided *in-the-moment* neural network performance integrity metrics (nPIMs) used to adjust the level of play required to properly engage long-term potentiation (LTP) and long term depreciation (LTD) network learning rules.

This study reports on t-test and Reliable Change Index (RCI) changes found within cognitive abilities’ performance metrics derived from the Woodcock Johnson Cognitive Abilities III Test. We compared pre and post scores from seven cognitive abilities considered dependent on executive cognitive control networks against seven non-executive control abilities. We observed significant improvements (*p* values 10 to 10^-22^) with large Cohen’s *d* effect sizes (0.78-1.20) across thirteen cognitive ability domains with a medium effect size (.49) on the remaining. The mean percent change for pooled trained domain was double that observed for pooled untrained domain, at 17.2% versus 8.3%, respectively. To further adjust for practice effects, practice effect RCI values were computed and further supported the effectiveness of the *r*CRT training (RCI-trained 1.4 - 4.8; untrained RCI 0.08-0.75).

## 1.0 Introduction

Mild to acute TBIs can lead to lingering changes in an individual’s neurologic performance, resulting in debilitating and far-reaching consequences in adaptive cognitive functioning. Annually, as many as 5.3 million people in the U.S. are thought to face challenges due to TBI-related disabilities [1]. However, the actual number of chronic TBI (> 6 months post-injury time) may be greater. This is due to limited testing sensitivity of typical testing methods for TBI based on conventional neuropsychological measures and/or conventional clinical imaging methods (e.g., CT, MRI scanning) coupled with a lack of public awareness with regard to mTBI symptoms [2,3].

Concussions represent 80% of the traumatic brain injuries (TBI) occurring each year in the United States [4-8]. Concussions are often related to sports injuries, but the bulk of concussions are due to motor vehicle accidents, falls, and situations involving sudden acceleration and/or deceleration of the head [4-8]. TBIs have been long considered an injury with little recourse, but recent awareness of the long-term effects of concussion has led to a renewed emphasis on treating TBI and concussions. If not treated properly, an instantaneous insult can be the beginning of a chronic disease process rather than just an isolated event. This disease process occurs across all levels of initial injury severity, from mild to severe [5-8]. For example, TBIs are implicated as a risk factor for cognitive impairments, reduced social functioning, psychiatric disorders, and chronic traumatic encephalopathy [9-13].

### TBI Cognitive Deficits

Cognitive deficits are some of the most common and undermining after-effects of a TBI. Such deficits may drastically interfere with an individual’s adaptive resiliency or ability to adapt to social and vocational collaborations, especially under mild stress conditions [14,15]. After a TBI, many major cognitive disruptions are triggered due to impaired gray matter or white-matter connections, often incurred by diffuse axonal injuries (DAI) [16]. DAIs foster multi-focal disturbances to axons that provide the structural basis of spatially distributed brain networks [17-21]. Consequently, a DAI often leads to interruptions in brain network connectivities, where these interruptions can be reflected in behavioral performance [17-21].In the context of rehabilitation, both active brain network performance and resting state functional connectivity (rsFC) metrics (measured using EEG or fMRI methods) are promising tools to measure neuroplasticity changes within an injured brain after injury [22-37]. These metrics can also provide evidence for experience-induced neuroplasticity changes acquired using *r*CRT methods [17-37].

Brain network performance deficits and/or DAI dysfunctions are often rooted in neural networks that sub-serve communications between larger networks. These larger networks support foundational neurobehaviors such as attention, memory, and executive functioning. White matter (WM) substructures of these networks and the efficiency of neural network hub connections (nodal connections within neural networks) demonstrate significant relationships with behavioral performance scores on intelligence testing [38-44]. Higher IQ scores correlate with higher nodal efficiency in the right anterior insula (AI) and dorsal anterior cingulate cortex (dACC), two hub regions within the salience network, with both regions shown to be vulnerable to mTBIs [38-44] and implicated in various mental health conditions [45-51]. Likewise, higher scores are linked to *lower nodal efficiency* in the left temporo-parietal junction area (TPJ).

Disruptions or lack of resiliency within these foundational neurobehaviors can impact various cognitive functions and emotional regulation abilities [17-21]. Spontaneous neural network reorganization resulting in a partial motor and cognitive recovery is commonly thought to occur in the first 3 to 6 months postinjury [52]. However, recent studies indicate that many deficits linger and are present years later [53-56]. Equally important, EEG performance studies indicate that the brain remodels or reorganizes to achieve a more normal behavioral performance; the remodeling may or may not have long term negative impact, depending on how the remodeling occurs [22-37]. Cognitive rehabilitation studies suggest that significant proper remodeling can be achieved by using cognitive rehabilitation exercises to reduce the cognitive and behavioral consequences of a mTBI [54-56]. Such exercises are the subject of this paper.

### Restorative Cognitive Rehabilitation Training (rCRT)

Cognitive rehabilitation training (CRT) methods are an organized, functionally oriented set of therapeutic activities based on a neural assessment. CRT treatments target the patient’s cognitive and behavioral deficits [54-56]. Fundamental to the CRT process is the brain’s ability to be remodeled through behavioral experience via neural plasticity changes, or the brain’s ability to reorganize and relearn, by redirecting maladaptive plasticity towards a more functional neural growth state [14-22]. CRT methods divide into restorative interventions (/CRT) and compensatory methods (cCRT). /CRT principally intervenes in cognitive disturbances or disrupted neural performance caused by brain impairment or disrupted function to promote brain performance normalization. cCRT seeks to establish alternative patterns of cognitive activity or create new patterns of movement through external support devices (e.g., adaptive aids, prosthesis) to improve the patient’s quality of life [54-56].

### rCRTRemediation Change Markers - Reliable Change Index, Cognitive Abilities and Resilience

Intelligence (cognitive ability) characterizes the ability to solve problems unrelated to previously learned knowledge, an essential element in resilient behavioural expressions [57]. These abilities underwrite encoding and use of new information with its efficient manipulation, representing a critical component of human cognition [39-45;58,59]. Equally, these abilities strongly predict educational and professional success [59], making the neural networks that support these operations obvious training targets. Cognitive neuroscience research supports this choice. For example, Santarnecchi et al. (2015) documented the association between individual intelligence quotients (IQ) and brain resilience by simulating targeted (specific network nodes) and random attacks using resting-state fMRI and graph analysis methods (n= 102 healthy individuals). The authors found enhanced brain resilience to targeted attacks (TA) were correlated with higher IQs in networks belonging to language and memory processing regions, whereas regions related to emotional processing were mostly supported in lower IQ individuals. These results suggest that both pre and post changes in IQ scores may be useful training predictors for recovery.

### Retrospective Chart Review Study

This study reviewed 200 clients who participated in a *r*CRT-based program to promote proper remodeling of neural function after a TBI. The approach employed the NeuroCoach® Training System, an automated rCRT activity/brain-computer interface system that develops targeted neural circuit responses towards resilient, flexible and adaptable behaviors. The approach applies algorithmically leveled brain training activities to support psychological resilience.

## 2.0 Methods

The study design employed a retrospective chart review to formulate results derived from participants who had previously participated in a BCI-augmented CRT program as a post-conventional treatment follow-on component of their mTBI recovery program. Our study protocol, 20-NEUR-101, was determined by an independent Institutional Review Board to be Exempt according to FDA 21 CFR 56.104 and 45CFR46.104(b)(4). Our use of data was retrospective, and data were processed for analysis in a manner which precluded identification of individuals. Informed consent was obtained in writing from all participants. To explore treatment effects, this study used a battery of Woodcock Johnson III (WJIIICA) assessments. Participant testing record results obtained from WJIIICA testing were structured with dependent pre- and post-test sampling using the same evaluation methods in both pre and post testing. Each participant received an individualized program designed to address neurobehavioral imbalances in their executive function and emotional regulation. Targeted treatment variables focused on remediating deficiencies observed in participants’ cognitive control, memory, attention, and executive function. Neurobehavioral imbalances were addressed using an advanced form of a CRT employing a BCI method to influence CRT training activities based on the cognitive information processing strength of each imbalance in real time [60-62]. Collection of data and subsequent analyses of those data were conducted by different persons, which helped both to ensure confidentiality and preclude bias from analysis.

### Participants

The TBI treatment group was composed of 200 participant records (n=200: 110 males and 90 females). The following training inclusion criteria were used: (1) mTBI derived from Sports, MVA, work related, and or recreational activity related; (2)>180 days post injury; (3) no histories of schizophrenia, bipolar disorder, eating or obsessive-compulsive disorder. Each group received the same pretest and posttest. Adult participants (aged 18 or older) previously classified with a closed head injury TBI were recruited from outpatient programs and private practices. The mean age of participants was 31.3 years. All records were de-identified using standard safe-harbor methods to protect the anonymity of individual health information. Participants volunteered for pre- and post-testing with treatment based on a deliberate self-selection convenient sample method. Volunteering did not affect the type of treatment received; specifically, those who did not volunteer or qualify for the study received the same BCI Amplified CRT Training as those who did. The treatment group was tested before treatment and upon treatment completion. All participants paid identical fees for treatment.

### Pre- and Post-Test Measures

This study employed neurophysiological performance, neurocognitive behavioral and psychometric measurements. The neurophysiological performance metrics were derived from resting and active state neuroelectric imaging methods, classic cognitive abilities task measures and from the Connor-Davidson Resilience Scale. Pre and post behavioral (classic task scores) and neural performance markers (age-normed Power Spectral Density (PSD) from resting state and event-related potentials) were obtained during the evaluation. Resting state neurometrics were derived from two FDA-registered databases (BrainDx, Neuroguide), using a z-score method to evaluate neurophysiological performance metrics. Active event-related potential neurometrics and z-score decision training metrics were obtained using a non-published proprietary database compiled from previous clinical and non-clinical cases (developed by the lead author). Access to the proprietary database for conducting neurometrics can be obtained by arrangment with the lead author at NTLGroup, Inc.

Figure 1 depicts the ten dependent (i.e., treatment) measures chosen from the Woodcock Johnson Cognitive Abilities III Assessment Battery (WJIII) [65] and five additional neurocognitive task measures derived from neurophysiological performance metrics. These measures were used to aid in *rCRT* exercise selection and in evaluating post training effectivness. The WJIII battery is a set of cognitive ability sub-tests based on the Cattell-Horn-Carroll (CHC) theory of cognitive abilities. The CHC theory provides a comprehensive framework for understanding the structure of cognitive information processing and the cognitive abilities required to support proper function. Five neurophysiological tasks were chosen to illuminate source-reconstructed neural network metric performance. These tasks included eyes-closed and eyes-open resting states, Flanker Task, Sternberg Working Memory Task and an Auditory Event Potential Task [66-72].

### Neurophysical/NeuroCognitive (NeuroCodex®) Pre-Post Evaluation

To obtain behavioral and temporal neural performance metrics, CHC tasks were presented to participants by the EventID task management program. Participants were seated in front of a computer screen and performed a battery of tasks derived from the WJIII battery. Each participant performed the cognitive battery while attached to a 19-channel EEG monitor (impedence below 5 kOhms) to record neuroelectric measures of EEG during the testing activities. After artifact rejection was performed on the EEG, behavioral and temporal neural metric measures were computed using classical ICA/PCA methods to obtain metrics for each test listed in Figure 1. The testing procedure began with a resting state eyes-closed and eyes-open condition as a baseline measure. Classical age-normed neurometrics were obtained based on standardized resting state quantitative EEG (qEEG) measures. These age-normed measures were included as baselines, compared against active ERP task measures as outlined in Figure 1.

To further support changes in resilient function, neural metric performance measures were obtained from five key source-reconstructed canonical networks that are considered to fine-tune behavior under variable environmental conditions. These networks are implicated in maintaining proper task performance and in general mental health preservation [67-73]. The program uses the Gordon et al [69] description of three distinct sets of connector hubs which integrate brain functional activités to model neurofunctional interactions. These three are Control-Default hubs, Cross-Control Connector hubs and Control-Prococessing hubs.

The five key networks include: *<u>Working Memory</u>* - the primary network that supports reasoning, expanded thought, and awareness by providing the mind a conscious workspace for information [69,70]; *<u>Cognitive Control Networks (CCN) -</u>* cognitive control incorporates processes involved in producing and preserving appropriate task goals, including suppressing irrelevant mental and physical activities that distract from achieving the desired set of task [69,70]; *CCN Subdivisions:* (1) *The Frontal-Parietal Network (FPN)* provides active online control, allowing it to adaptively initiate and adjust control [69,70]; (2) the *Cingulate-Opercular Network (CON)* provides stable ‘set-maintenance’ (state maintenance) over the entire task epoch or behavioral strategy[69,70]; (3) the *Salience Network (SN*, the *Attention Networks plus Insula Network)* is involved in rapid detection of goal-relevant events and facilitation of access to appropriate cognitive resources by interacting with multiple functional systems, thereby supporting a wide range of cognitive processes[69,70]. The *<u>Default Mode Network (DMN)</u>* is implicated in the brain’s default resting state conditions and in its ability to sustain task performance. The DMN is composed of functionally specialized subsystems, with the anterior DMN (i.e., medial Prefrontal Cortex (PFC)) associated with identifying stimuli as self-salient, whereas the posterior DMN region (with the parahippocampal gyrus) is involved in autobiographical search and memory retrieval. Mechanisms within the DMN are implicated in regulating emotional reactivity and may play a key role in the empathic process by establishing a distinction between other- and self-related feelings [69-73]. Regarding congruent cognitive/behavioral health performance, a close relationship exists between empathy and executive regulatory mechanisms. Sluggish and/or poor (dis)engagement of the DMN is a noted biomarker within several mental health conditions, including depression and attention deficit disorders [69,73]. The opposing relationship between DMN and cognitive control networks may influence the ability to exert cognitive control [69,73] and play an important role in the regulation of mind-wandering and rumination that impacts task performance [73].

**Fig 1.**
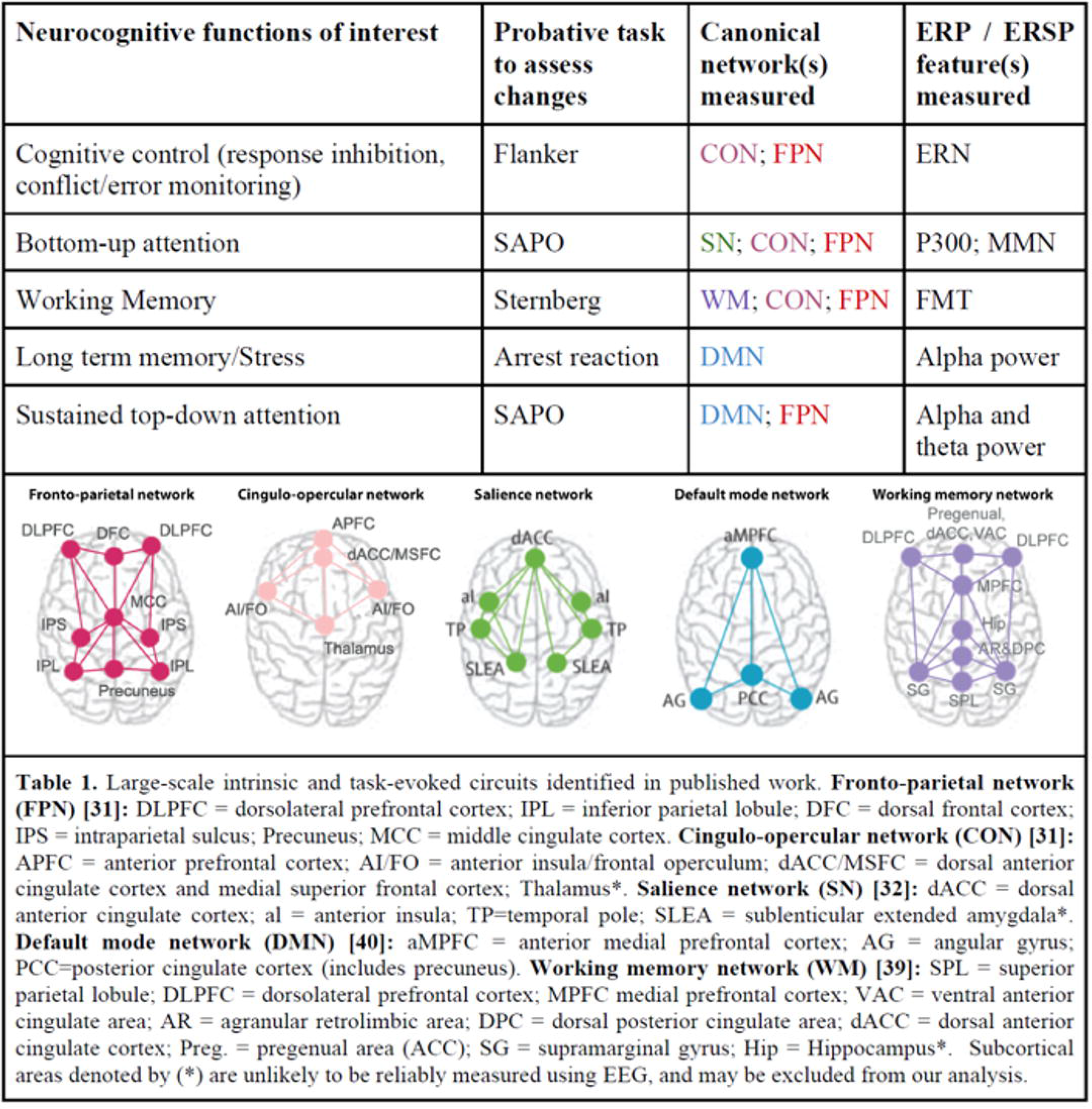
Neurocognitive Functions of Interest and Canonical Networks

### Training Procedure

Immediately after initial evaluation, participants used the NeuroCoach® Training System three times per week for twelve weeks (approximately 30-40 minutes per session); participants were then reassessed. All participants completed a non-verbal cognitive enhancement/neuro-remodeling treatment program that monitors and evaluates a user’s defined neural networks system performance status in real-time. Between 48 and 80 sessions of extensive training (approximately 30-40 minutes per session) were completed before final re-evaluations were completed. The training system is rooted in modern *r*CRT methods, incorporating a neural network BCI monitoring interface. The BCI provides neural network performance integrity metrics (nPIMs), originated from one or more of the three control connector hub systems. The nPIMs inform the leveling training algorithm as it adjusts program training intensity levels. The BCI adjusts the difficulty level for each training activity based on *in-the-moment* brain performance metrics. Individual nPIMs are derived from the neural network systems that support various cognitive functions being trained and are user-selectable. The *r*CRT methodology is implemented through a selectable set of computer activities specific to individual needs and engages the desired brain network systems and cognitive functions. Each activity is based on classic neuroscience paradigms [60-69]. The BCI interface informs the trainer, the user, the *r*CRT activity in real time the current neural network performance integrity status based on the user’s present nPIMs state.

Each *r*CRT activity incorporates a performance leveling algorithm (PLA) to adjust the intensity of the activity by rendering the pursuit to be either more or less intense. Unique in our method is that the PLA encompasses both nPIMs and behavioral responses (e.g., response times, accuracy) to adjust the level of intensity of the activity. This adjustment is based on the real-time performance ability of the user, and targets the intensity required to properly engage long-term potentiation (LTP) and long-term depreciation (LTD) network learning rules [75-83]. Difficulty is adjusted based upon current responses, with the goal of a proper ratio of neurocircuit engagement as opposed to a certain level of correct responses. The intention of the performance-leveling algorithm is to adjust the level of pursuit play to a comfortable level, allowing the user to progress through the activity successfully while simultaneously focusing on developing and/or strengthening the performance integrity of the neural system being trained.

### NeuroCoach® Training Module Example and Description

*The Split-Attention* application (a NeuroCoach® training module) is an adaptive process-based, nonverbal training technique designed to aid in re-setting/enhancing the attention (ATN), working memory (WMN), frontal parietal (FPN) and salience networks (SN). Split-Attention uses a relaxation and restorative framework that allows the trained networks to regain or obtain a natural homeostatic balance needed to maintain a desired level of performance as it drives the user towards increased capacity, neural efficiency and performance resilience. Neurobehaviorally, the application focuses on training the useful field of view (visual attention), working memory, cognitive speed, task switching, and multiple attention abilities, all in one application.

The lead author has used this application clinically for ten years with brain-injured and learning-disabled populations. The application promotes a relaxed sustained attentional focus in professional athletes and supports restorative cognitive enhancement. The Split-Attention exercise satisfies The Institute of Medicine’s Checklist criteria for brain training [39].

### Analysis of Pre- and Post-rCRT Scores

Scoring data for all participants were pooled within a Pandas data frame and analyzed as a batch for means, standard deviations and pre/post *r*CRT changes by means of a Python script. P-values comparing pre/post *r*CRT subject scores were calculated from entire population distributions by using the two-sided Kolmogorov-Smirnov algorithm within SciPy. This method consistently produced more conservative (i.e., larger) p-values than other methods which consider only population means and standard deviations.

### Analysis of Practice Effects - RCI Calculations

The RCI technique used to correct for practice effects and measurement error is defined as ((*X*_2_−*X*_1_ − (*M*_2_ − *M*_1_))/SDD [74,75] where *X*_1_ is the measured pretest score, *X*_2_ the post-test score, SDD the standard deviation of the group test-retest difference, *M*_1_ the control group mean pretest score, and *X*_2_ the control group mean post-test score. As a retrospective chart review, the study did not use control subjects and therefore obtaining measures of *M*_1_ and *M*_2_ are not directly available. However, several studies have determined that the estimated change in cognitive test retest scores range between .25 and .33 of a typical standard deviation [74-76]. Applied to standard scores, *M*_1_ *M*_2_ values would range between 3.75 and 5.0. Table 2 presents results for the 14 Woodcock Johnson Variables and the RCI results from all tests.

**Table 1.** Summary of the Pre- and Post-299 treatment Training Results Large-scale intrinsic and task-evoked circuits identified in published work. **Fronto-parietal network (FPN) [31]**: DLPFC = dorsolateral prefrontal cortex: IPL = inferior parietal lobule: DFC = dorsal frontal cortex: IPS = intraparietal sulcus: Precuneus; MCC = middle cingulate cortex. **Cingulo-opercular network (CON) [31]**: APFC = anterior prefrontal cortex: AIFO = anterior insula frontal operculum: dACC/MSFC = dorsal anterior cingulate cortex and medial superior frontal cortex; Thalamus*. **Salience network (SN) [32]**: dACC = dorsal anterior cingulate cortex; al = anterior insula; TP=temporal pole: SLEA = sublenticular extended amygdala*. **Default mode network (DMN) [40]:** aMPFC = anterior medial prefrontal cortex; AG = angular gyrus: PCC=posterior cingulate cortex (includes precuneus). **Working memory network (WM) [39]**: SPL = superior parietal lobule: DLPFC = dorsolateral prefrontal cortex; MPFC medial prefrontal cortex: VAC = ventral anterior cingulate area; AR = agranular retrolimbic area: DPC = dorsal posterior cingulate area; dACC = dorsal anterior cingulate cortex; Preg. = pregenual area (ACC); SG = supramarginal gyrus; Hip = Hippocampus*. Subcortical areas denoted by (*) are unlikely to be reliably measured using EEG, and may be excluded from our analysis.

| **<u>Trained</u>** | **Pre-Mean**<br>**(SD)** | **Post-Mean**<br>**(SD)** | **Change**<br>**(SD)** | **t Stat** | **$P_{2tail}$** | **Cohen's**<br>**$d$** | **RCI**<br>**pe3.75** | **RCI**<br>**pe5.00** |
| --- | --- | --- | --- | --- | --- | --- | --- | --- |
| *General Intellectual Ability* | 100.35<br>(12.73) | 115.64<br>(12.78) | 15.29<br>(7.67) | 28.13 | <.01 | 1.2 | 3.14 | 2.8 |
| *Thinking Efficiency* | 100.05<br>(15.32) | 113.42<br>(14.89) | 13.37<br>(10.03) | 18.86 | <.01 | 1.04 | 1.92 | 1.67 |
| *Concept Formation* | 102.42<br>(15.74) | 113.17<br>(12.16) | 10.75<br>(7.79) | 13.41 | <.01 | 1.18 | 1.36 | 1.12 |
| *Working Memory* | 102.08<br>(17.49) | 117.21<br>(17.80) | 15.13<br>(12.64) | 16.94 | <.01 | 1.04 | 1.8 | 1.67 |
| *Numbers Reversed* | 101.46<br>(19.60) | 119.54<br>(19.01) | 18.08<br>(15.20) | 15.78 | <.01 | 1.13 | 1.88 | 1.72 |
| *Visual Auditory Learning* | 95.63<br>(18.52) | 113.96<br>(19.05) | 25.24<br>(25.31) | 16.91 | <.01 | 1.21 | 2.23 | 2.03 |
| *Visual Audio Delayed* | 74.4<br>(33.62) | 103.65<br>(31.81) | 29.25<br>(15.32) | 16.91 | <.01 | 1.11 | 4.83 | 4.6 |
| **<u>UnTrained</u>** | **Pre-Mean**<br>**(SD)** | **Post-Mean**<br>**(SD)** | **Change**<br>**(SD)** | **t Stat** | **$P_{2tail}$** | **Cohen's**<br>**$d$** | **RCI**<br>**pe3.75** | **RCI**<br>**pe5.00** |
| *Verbal Ability* | 97.62<br>(9.74) | 105.135<br>(11.05) | 7.52<br>(7.02) | 15.13 | <.01 | 0.76 | 0.63 | 0.42 |
| *Phonemic Awareness* | 104.93<br>(13.00) | 114.83<br>(13.53) | 9.9<br>(8.74) | 16.01 | <.01 | 0.87 | 0.66 | 0.52 |
| *Verbal Comprehension* | 97.59<br>(9.76) | 105.25<br>(11.08) | 7.66<br>(7.33) | 14.78 | <.01 | 0.85 | 0.65 | 0.44 |
| *Incomplete Words* | 101.16<br>(18.00) | 113.14<br>(18.94) | 11.99<br>(14.27) | 11.88 | <.01 | 0.92 | 0.88 | 0.75 |
| *Sound Blending* | 106.28<br>(11.82) | 113.47<br>(11.31) | 7.19<br>(7.90) | 12.87 | <.01 | 0.78 | 0.48 | 0.31 |
| *Spatial Relations* | 103.70<br>(12.74) | 112.22<br>(11.38) | 8.52<br>(9.78) | 12.32 | <.01 | 1 | 0.52 | 0.38 |
| *Visual Matching* | 98.73<br>(12.12) | 104.22<br>(12.88) | 5.49<br>(8.06) | 9.64 | <.01 | 0.49 | 0.27 | 0.08 |

## 3.0 Results

Subjects completed identical Woodcock Johnson III assessments before and after *r*CRT treatment. The battery included fourteen assessments in the following areas: General Intellectual Ability (GIA), Thinking Efficiency, Concept Formation, Working Memory, Numbers Reversed, Visual Auditory Learning, Visual Audio Delayed, Verbal Ability, Phonemic Awareness, Verbal Comprehension, Incomplete Words, Sound Blending, Spatial Relations and Visual Matching. *r*CRT treatment explicitly targeted development in neurocircuits related to the first seven of these areas but did not explicitly target development in the second seven areas. Given the absence of a control group in this retrospective chart review study, measuring performance in both targeted and untargeted areas provided some assessment of the magnitude of specific treatment effects.

Table 2 summarizes the pre- and post-treatment results across all Woodcock Johnson III assessments. Histograms of pre- and post-training scores for individual areas are available in Supplementary Materials (Figures S1-S14). Notably, we observed significant improvements (p values ranged from 10^-4^ to 10^-22^) across all Woodcock Johnson III areas, as might be expected after many sessions of intensive rCRT. To assess the differential impact of explicitly targeting an area within the *r*CRT program, we adjusted these observed improvements to reflect the percent change within each area, and compared the pooled percent changes observed in trained areas against those observed for untrained areas. Figure 1 displays histograms for these pooled changes. The mean percent change for trained areas was double that observed for untrained areas, at 17.2% versus 8.3%, respectively. Figure 2 shows percent changes observed across each area, ranked by magnitude, and highlights how consistently trained areas received a greater percent change than those observed for untrained areas.

We observed no significant correlation in pre-training, post-training or change scores as a function of subject sex or age, regardless of the area assessed (Supplementary Materials, Figures S1-S14).

**Fig 2.**
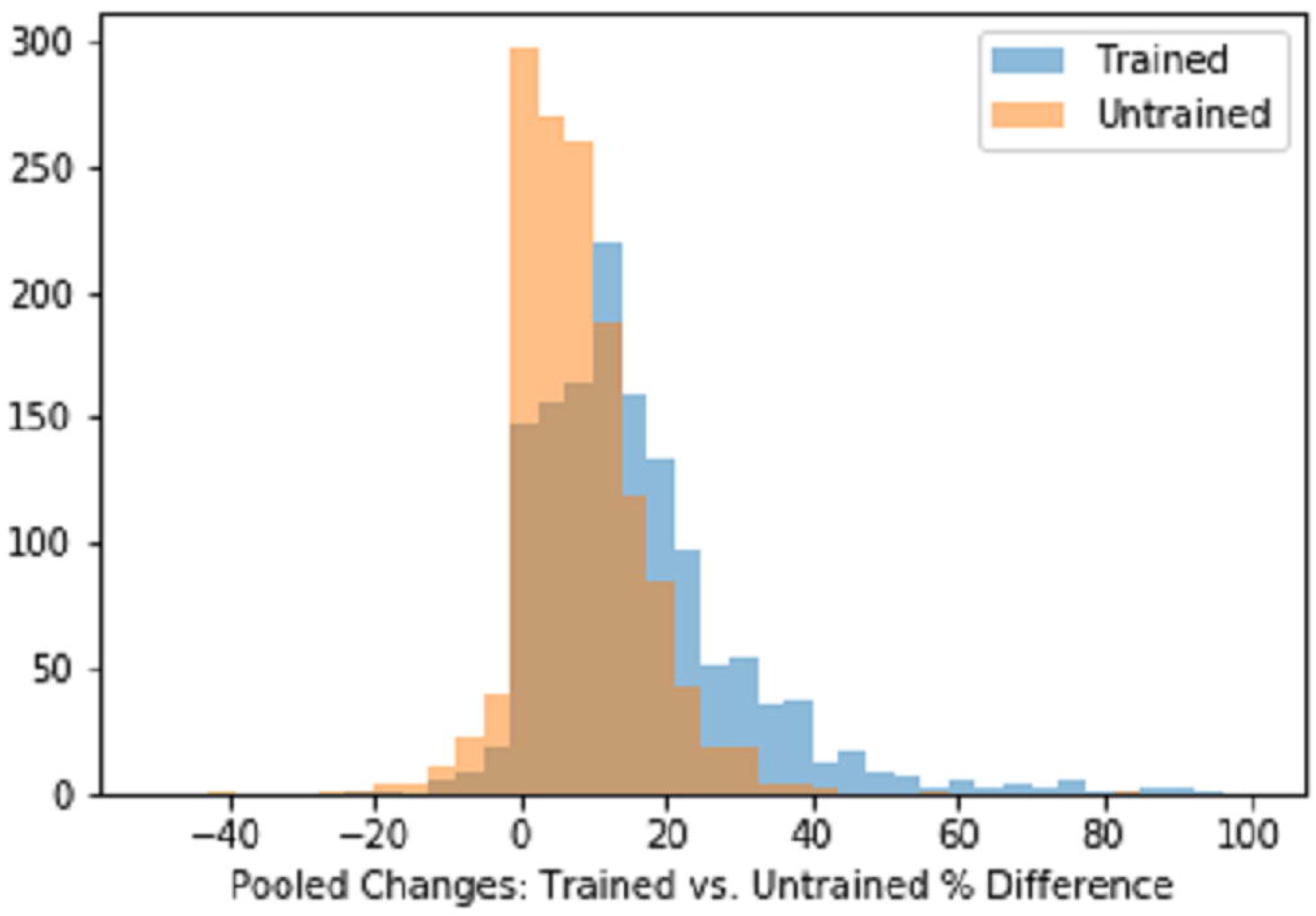
Histogram of Pooled Changes

**Fig 3.**
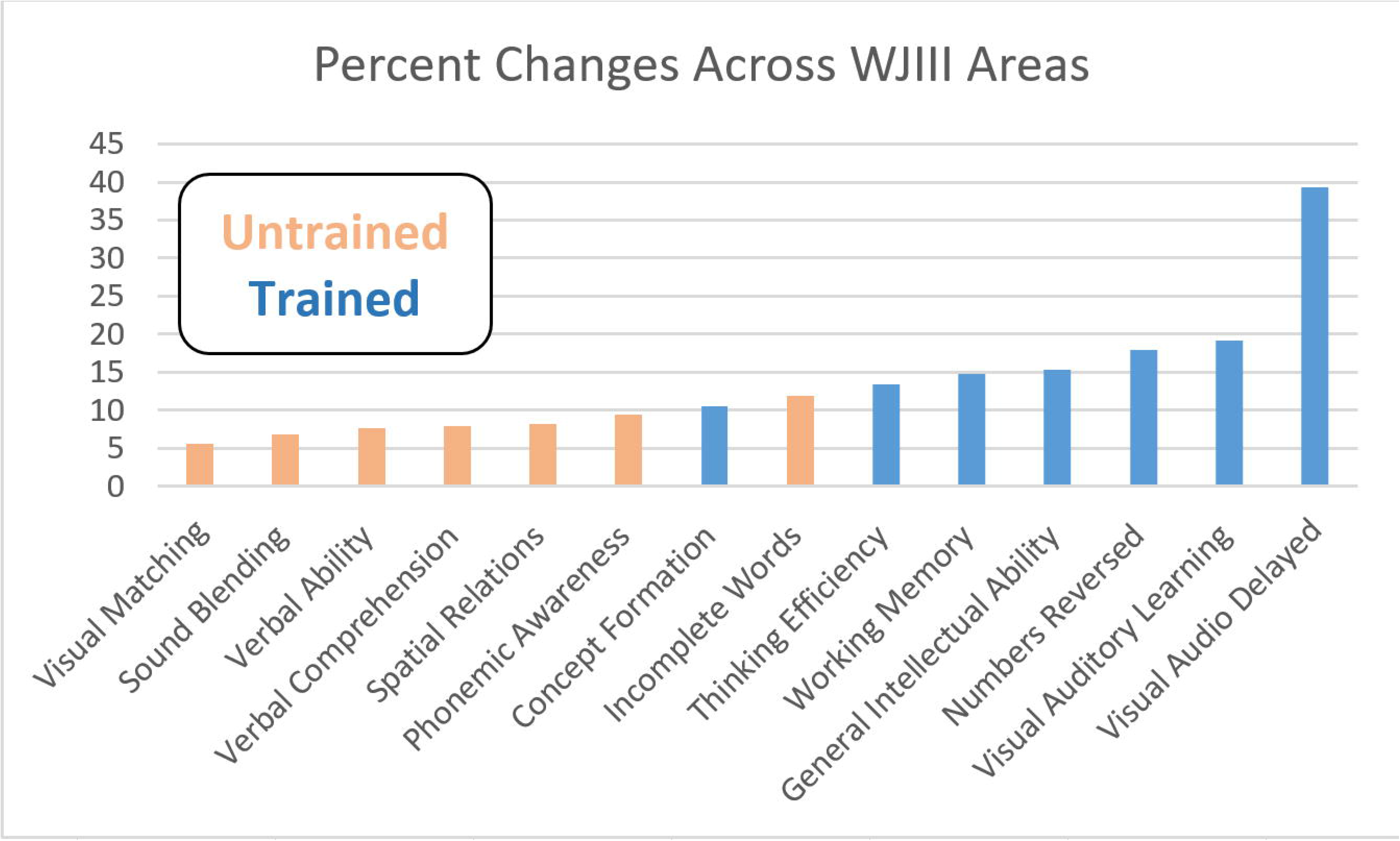
Percent Changes Observed Across each Area

## 4.0 Discussion

Previous mTBI CRT program meta-analyses report medium effect size training improvements [75-77]. As such, the study authors recommend CRT methods as a viable method for treating mTBIs [75-77]. Traditional mTBI treatment programs generally begin with neuropsychological behavioral testing that do not include a neuroimaging exam. As a result, training is focused on behavioral deficits found within attentional, memory, or other executive domains, without considering possible neural network performance interruptions nor possible reduced neural integrative effects. In contrast, this study explored possible neural network performance interruptions by choosing training targets based on possible neural network performance interruptions based on a standardized cognitive task-based neuroimaging exam. Additionally, to support optimal network remodeling during /CRT training, each training activity was guided by network performance nPIMs provided by a BCI interface in support of proper neural functional remodeling needed to support resilient, flexible and adaptable behaviors. More expressly, the /CRT training program focused on improving neural network functional performance to support long-term potentiation (LTP) and long-term depreciation (LTD) network learning rules during the training process.

Group level t-test and practice effect RCI value changes support significant positive changes within important cognitive abilities’ performance metrics known to support executive cognitive control abilities needed in resilient, flexible and adaptable behavioral expressions. /CRT training target selection focused on cognitive control training activities. We anticipated a positive training effect to occur in the all measured cognitive domains due to general cognitive improvement in cognitive network efficiency. However, we further expected a greater improvement in the executive function metrics due to the focus in the training. Pre and post scores from seven cognitive abilities considered dependent on executive cognitive control networks were compared against seven non-executive control abilities and supported our expectations. We observed significant improvements *(p* values 10^-4^ to 10^-22^) with large Cohen’s *d* effect sizes (.78-1.20) across thirteen cognitive ability domains with a medium effect size (.49) on the remaining. The mean percent change for pooled trained domain was double that observed for pooled untrained domain, at 17.2% versus 8.3%, respectively. To further adjust for practice effects, practice effect RCI values (based upon literature known adjustments) were computed and further supported the effectiveness of the /CRT training (RCI-trained 1.4 - 4.8; untrained RCI .08-.75) on the executive control networks.

In summary, this mTBI study demonstrates a strong possible increase in training effects over conventional *r*CRT methods. This was achieved by first using a neuroelectric imaging exam based on EEG source reconstructed neural network methods used in *r*CRT training target selection. Second, we augmented individual *r*CRT activities with a BCI interface to monitor and compute *in-the-moment* neural network performance integrity metrics (nPIMs) needed to align the level of activity engagement. Activity level computations were required to properly manage cognitive loads and to properly engage long-term potentiation (LTP) and long-term depreciation (LTD) network learning rules. From our experience, this automated approach to classical *r*CRT methods offers two extensions over traditional pen and pencil, or computer game CRT approaches: 1) tailoring the selection of the *r*CRT training procedures based on neural network performance metrics derived from EEG source reconstruction neuroelectric imaging evaluations to isolate underlying neural network disruptions; 2) amplifying neural network regional training by means of BCI treatment amplification.

In general, the training program assumes that coupling key, resilience-supporting neural circuits with proper problem-solving skills promotes the emergence of resilient, adaptive behaviors. Based upon program participant subjective reports, we found that in the context of daily living, this emergence means proper brain based behavioral health expressions that allow return to productive work, social reintegration and improvement in one’s quality of life. In other contexts, such as in sports, this emergence means increased sports performance for both injured and non-injured athletes.

This retrospective chart review was limited by the lack of a control group, although comparing explicitly trained versus untrained cognitive areas provided some measure of the effect of treatment. Future work will further “mine” retrospective data to inform the design and focus of controlled, prospective studies. In addition, customized individual *r*CRT programs will benefit from the insights gleaned from analysis of our database of retrospective data.

## Data Availability

Raw qEEG and cognitive testing data cannot be shared publicly because of privacy issues. De-identified pre/post cognitive testing data which underlie the results of the study are available from Curtis Cripe, NTL Group, Inc., 602-799-2051

